# Genetic Determinants of Disease Trajectory in Sarcoidosis

**DOI:** 10.1101/2024.12.29.24319750

**Authors:** Soumeen Jin, Susanna Kullberg, Pernilla Darlington, Tomoko Iseda, Olga Chuquimia, Vasiliki Vasila, Ingrid Kockum, Anna Smed-Sörensen, Anders Eklund, David Ellinghaus, Antje Prasse, Stefan Schreiber, Joachim Müller-Quernheim, Kristen R Mathias, Mehdi Mirsaeidi, Leonid Padyukov, Paolo Spagnolo, Michelle Sharp, Natalia V. Rivera

**Author notes:** Correspondence should be addressed to: Natalia V Rivera, PhD, Division of Immunology and Respiratory Medicine, Department of Medicine Solna, Karolinska Institutet, Karolinska University Hospital, Solna, and Center for Molecular Medicine, Karolinska University Hospital, Stockholm, Sweden., Division of Pulmonary & Critical Care Medicine, Johns Hopkins University School of Medicine, Baltimore, Maryland, USA.

## Abstract

**Background:** Sarcoidosis is a complex granulomatous, systemic inflammatory condition of unknown origin, characterized by diverse clinical profiles, most notably Löfgren’s syndrome (LS) and non-LS sarcoidosis. We investigated the genetic underpinnings of disease trajectory defined by chronic and nonchronic phenotypes.

**Methods:** Two pulmonologists assessed sarcoidosis disease trajectory over two years and classified patients as chronic or nonchronic. We conducted genetic analyses of disease trajectory in a Swedish cohort comprising 1,484 sarcoidosis cases (775 chronic and 709 nonchronic) and 5,817 controls, for a total of 7,301 individuals. We further stratified genetic analyses by LS and non-LS and performed SNP-heritability analyses, gene-based analyses, eQTL enrichment, and pathway analyses to assess the biological significance of the identified genomic loci.

**Results:** Genetic associations with chronic and nonchronic sarcoidosis identified distinct loci. In chronic sarcoidosis, the lead signal was rs3135387 (OR = 1.659, 95% CI 1.436 - 1.916) near *HLA-DRA*. In nonchronic sarcoidosis, the strongest signal resided within *TSBP1* rs2849018 (OR = 3.70, 95% CI 3.14 - 4.37). Notable clinical-subtype signals included rs2849018 in LS nonchronic sarcoidosis (OR = 5.90, 95% CI 4.83–7.22), rs3117224 in non-LS chronic sarcoidosis (OR = 1.683, 95% CI 1.439 - 1.968), and rs59344869 in non-LS nonchronic sarcoidosis (OR = 2.424, 95% CI 1.738 - 3.380). SNP heritability was 32.8% (95% CI 23.9–41.7%) for chronic and 47.5% (95% CI 37.5–57.6%) for nonchronic sarcoidosis.

**Conclusions:** This study highlights the distinct genomic features that differentiate disease trajectories across sarcoidosis clinical phenotypes, underscoring the critical involvement of immune-related genes and regulatory networks. These findings pave the way for further molecular studies and patient stratification based on genetic profiles.

## Introduction

Sarcoidosis is a systemic granulomatous inflammatory disease that can impact any organ, most commonly involving the lungs and lymph nodes. Its clinical disease trajectory varies widely, affecting multiple organs simultaneously. The disease generally falls into two main categories: resolving forms, like Löfgren’s syndrome (LS), and non-resolving forms, such as non-Löfgren’s sarcoidosis, which may lead to pulmonary fibrosis [1]. Recent research emphasizes the need for more detailed classification of clinical phenotypes, reflecting the disease’s complexity and diversity [2–4].

Although more than 50-60% of patients experience spontaneous resolution or remission, approximately 40-50% of individuals develop a clinically progressive course often complicated by the development of pulmonary fibrosis and organ failure [5–7]. Despite this significant finding, the underlying mechanisms driving disease trajectory in sarcoidosis remain largely unexplored. Furthermore, the absence of standardized characterization of disease trajectory, coupled with the lack of a definitive biomarker to forecast clinical outcomes, impedes clinicians in predicting patient prognoses. Gaining insight into the genetic architecture of disease trajectory is an essential step toward achieving personalized medicine objectives and enhancing care for individuals with sarcoidosis.

Accumulating research suggests that sarcoidosis results from a complex interplay between genetic factors and environmental triggers [8–10], particularly in individuals carrying predisposing alleles [11].

Sarcoidosis manifests variably among individuals with a genetic predisposition, with environmental triggers, occupational exposures, and lifestyle behaviors modifying disease risk and its etiology and disease trajectory [12, 13]. As immunological and genetic understanding of sarcoidosis advances, growing evidence suggests that sarcoidosis is immunologically driven, with overlapping immunological cellular profiles, immune dysregulation in response to an unknown antigen, and genetic associations [14, 15]. Of note, proinflammatory cytokines were expressed 10 years before disease onset, supporting the theory of an immune-mediated susceptibility in a specific group of sarcoidosis patients [16]. Over the past two decades, class II genes in the human leukocyte antigen (HLA) region have been linked to disease trajectory, clinical outcomes, and target-organ phenotypes in sarcoidosis [17]. For instance, patients with Löfgren’s syndrome (LS) typically have a favorable prognosis, especially carriers of *HLA-DRB1*03:01* (DR3) allele (also referred to as DR3+). In DR3+ LS, pulmonary CD4+ T cells show a striking accumulation of T cell receptors that use the TRAV12-1 alpha chain (formerly AV2S3) and are associated with resolving disease [18]. In contrast, patients with non-LS often present with a gradual onset and a range of clinical features, tending to progress toward a chronic form of the disease, which is associated with a poorer prognosis and organ failure [19, 20].

Similar to LS, HLA genes play a crucial role in target organ phenotypes and overall disease trajectory, also in non-LS [21].

*HLA-DRB1* alleles have different biological roles in autoimmune diseases, disease outcomes, and chronicity across populations [22]. In sarcoidosis patients of Swedish ancestry, *HLA-DRB1 *07, *14,* and *\*15* alleles often have a chronic disease course, while *HLA-DRB1 *01* and *03 alleles tend to confer protection against persistent disease [22]. Conversely, *HLA-DRB1*11:01* has been linked to an increased risk of disease in both European and African American populations [23].

This study investigated whether different genetic structures affect disease trajectory in sarcoidosis, aiming to define chronic and nonchronic phenotypes and assess genetic effects after stratifying clinical phenotypes as LS and nonLS.

## Materials and Methods

### Study population

Two pulmonary specialists examined a Swedish sarcoidosis cohort to analyze disease trajectory phenotypes using chronic and nonchronic definitions. We further characterized the cohort using these definitions and stratified it by clinical phenotype, including LS and non-LS. We analyzed data from our local registry at Karolinska University Hospital, Stockholm, Sweden, which includes clinical data collected from 1987 to 2024. The Stockholm Region Review Board approved the study (DNR: 2024-03916-02). All participants provided written informed consent and permitted the use of their DNA for research purposes. We collected blood samples and baseline characteristics at study entry. Briefly, we enrolled patients for investigation at the Sarcoidosis Center, Karolinska University Hospital, Solna, Sweden. The diagnosis was made using the criteria outlined by the World Association of Sarcoidosis and Other Granulomatous Disorders (WASOG, Statement on Sarcoidosis, 1999). For this study, 1,484 sarcoidosis patients (709 nonchronic and 775 chronic) for disease trajectory between 2014 and 2024 were included. We determined disease trajectory phenotypes at the two-year follow-up. Healthy controls included 5,817 individuals recruited from two large-scale epidemiological cohorts, the Environmental Investigation of Rheumatoid Arthritis (EIRA) [24] and the Epidemiological Investigation of Risk Factors for Multiple Sclerosis (EIMS) [25].

### Phenotype definition of disease trajectory

We assessed disease trajectory at a two-year follow-up in 1,484 patients. We defined a nonchronic case as normalized radiography and pulmonary function, no symptoms, normal biochemical parameters, and no signs of extrapulmonary disease two years after diagnosis. Patients with only residual calcified lymph nodes and no signs of active disease two years after diagnosis were also defined as having a nonchronic disease. Chronic disease was defined as persistence or progression of chest radiographic changes and/or decreased/deteriorating lung function and/or clinical signs of disease activity, with symptoms and/or biochemical evidence of disease activity and/or extrapulmonary sarcoidosis, at 2 years after diagnosis.

#### Genotyping

We genotyped Swedish sarcoidosis patients at the SNP&SEQ Technology Platform at Uppsala University, Sweden, using an Illumina GSA-MD single-nucleotide polymorphism (SNP) array with ∼700,000 markers. We obtained healthy controls (HC) from the EIRA and EIMS [23] cohorts, which we also genotyped on the same platform.

#### Quality control

We performed quality control (QC) and filtering separately for each cohort (or batch) using PLINK 1.9b [26]. Using PLINK binary files (bed, bim, and fam), we extracted the autosomal chromosomes (1-22) for QC analyses. Individuals with a genotype call rate <97%. SNPs with minor allele frequency (MAF) <1%, SNPs with Hardy-Weinberg equilibrium (HWE) P < 1 × 10⁻⁷ in controls, and individuals with a relatedness coefficient (F ≥ 0.125) were excluded. To account for ancestry, we calculated principal components in PLINK v1.9b using 38,027 linkage-disequilibrium-pruned typed SNPs and included the first two principal components (PC1 and PC2) as covariates.[27]

#### Imputation

SNP imputation was conducted on the Michigan Imputation Server[28] using the European Reference Panel (build GRCh37/hg19) available in the Haplotype Reference Consortium (HRC)[29]. Quality control on imputed SNPs included removal of SNPs with MAF < 1%, imputation quality (R² < 0.3), and a missing genotype rate > 3%. Post-imputation QC resulted in 4,093,469 SNPs and 7,001 individuals (1,484 cases and 5,817 controls).

### Statistical analysis

We calculated descriptive statistics (mean, median, standard deviation) in R. We assessed count data distributions using Fisher’s exact test or Pearson’s Chi-squared test with Yates’ continuity correction, as appropriate, as implemented in R.

#### *HLA-DRB1* analysis

We evaluated the distribution of *HLA-DRB1* alleles across the disease trajectory groups (chronic and nonchronic) using a chi-square test. To account for multiple testing, we applied False Discovery Rate (FDR) correction on *HLA-DRB1* alleles and considered results significant at an FDR < 0.05. We also examined the distribution of *HLA-DRB1* alleles in clinical phenotypes LS and nonLS. Furthermore, we assessed how *HLA-DRB1* alleles influence disease trajectory groups and clinical phenotypes using logistic regression with FDR correction for multiple comparisons.

##### Association test

We tested genetic associations using an additive model in logistic regression comparing sarcoidosis clinical phenotype with healthy controls. The regression model regressed the binary outcome on SNPs, adjusted for sex, age, and the first two principal components (PCs). We performed principal component analyses on pruned genotypes (38,027 SNPs) using PLINK v1.9b [26]. SNP pruning was calculated using a linkage disequilibrium (LD)-based function (r² = 0.25) implemented in PLINK.

#### SNP Heritability

We estimated observed-scale SNP heritability (h^2^) using univariate single-component genomic restricted maximum likelihood (GREML), implemented in Genome-wide Complex Trait Analysis (GCTA) v1.95.3, among unrelated individuals, with sex, PC1, and PC2 included as covariates [29]. To transform to liability-scale heritability (h2L), we derived population prevalence estimates for chronic and nonchronic sarcoidosis and their sub-phenotypes from epidemiological evidence [29, 30]. These references yielded population prevalence estimates of 45.60, 114.40, 7.68, 37.92, 40.32, and 74.08 per 100,000 for chronic, nonchronic, LS chronic, non-LS chronic, LS nonchronic, and non-LS nonchronic sarcoidosis, respectively. Observed-scale estimates were transformed using the equation for ascertainment-corrected liability-threshold transformation defined by Lee et al. [30]

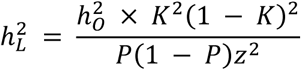

where K denotes the assumed population prevalence, P is the case proportion in the analyzed sample, t = Φ⁻¹(1 − K), and z = φ(t). Given the available sample size, we used single-component GREML as suggested by Visscher et al. [31]. The final analysis sample sizes were chronic (n = 3,414), nonchronic (n = 3,405), LS chronic (n = 2,957), non-LS chronic (n = 3,363), LS nonchronic (n = 3,222), and non-LS nonchronic (n = 3,089).

##### Gene-based analyses

To identify relevant genes, we used Multi-marker Analysis of GenoMic Annotation (MAGMA) [32] and applied a Bonferroni threshold of P < 2.687 × 10⁻⁶ (0.05/18,605 tested genes). The denominator was the 18,605 genes present in each supplied MAGMA output.

##### Enrichment, functional mapping, and annotation

We performed post-GWAS using Functional Mapping and Annotation of Genome-Wide Association Studies (FUMA-GWAS) [33]. Given the tissue and cell-type relevance to sarcoidosis, we focused on eQTLs in the lung, whole blood, and immune cells.

##### Pathway analyses

We conducted pathway analyses using the QIAGEN Ingenuity Pathway Analysis (IPA) Knowledge Base, primarily using Core Analysis of MAGMA gene-level results (P < 0.05) for chronic and nonchronic, and for clinical phenotypes LS and non-LS [34, 35]. The top 15 pathways were visualized by merging the overlapping pathways in each chronic–nonchronic pair and ranked according to

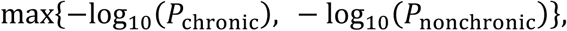

in either chronic or nonchronic phenotype across overall, LS, and nonLS sarcoidosis.

## Results

Table 1 shows baseline characteristics of the Swedish cohort used to characterize disease trajectory. We characterized disease trajectory in 1,484 patients (709 nonchronic and 775 chronic) between 2014 and 2024. Figure 1 shows the cohort distribution by disease trajectory and sarcoidosis clinical phenotypes.

**Figure 1.**
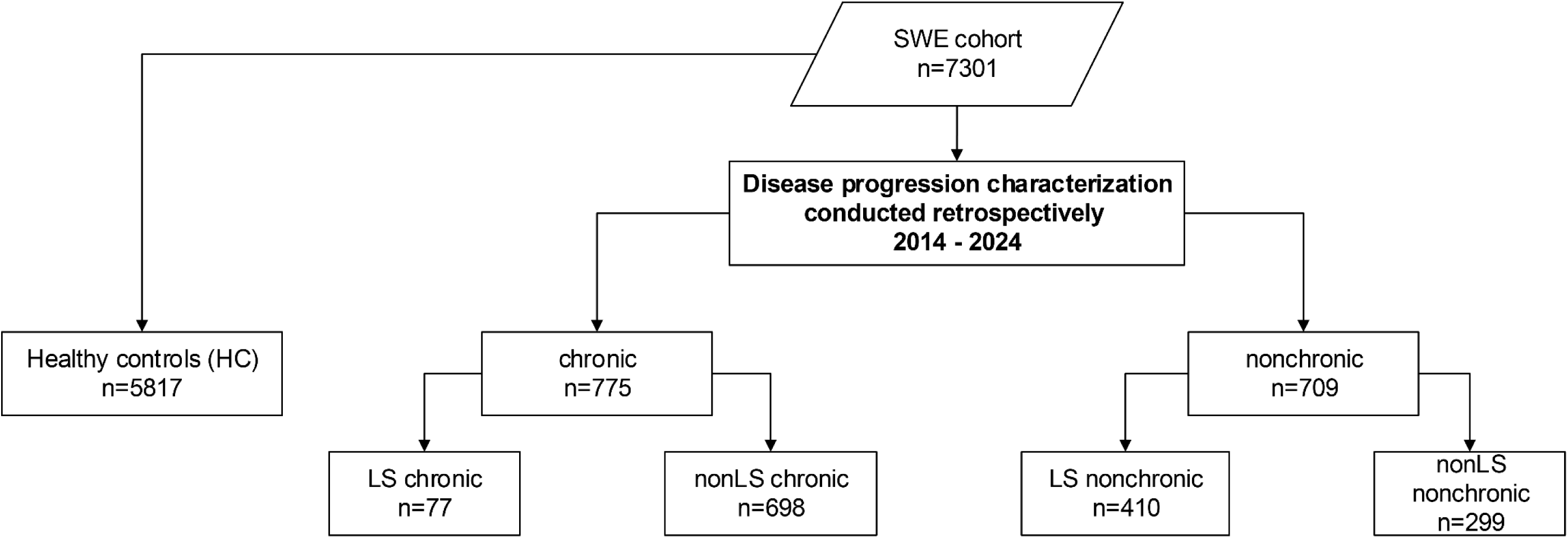
Swedish cohort characterization and study design

**Table 1.** Cohort characteristics.

|  | Chronic<br>(n = 775) | Nonchronic<br>(n= 709) | P-value | Healthy controls<br>(n = 5,817) |
|---|---|---|---|---|
| **Biological sex** |  |  |  |  |
| % males | 56.3% | 43.7% | 1.08e-04 |  |
| % females | 45.8% | 54.2% |  |  |
| **Age (years)** (Mean ± SD) | 45.5 ± 11.8 | 41.6 ± 11.9 | 2.66e-13 | 55 ± 11.7 |
| **Chest X-ray stage**<br>0/1/2<br>3/4 | 27/201/374<br>116/57 | 25/393/256<br>35/0 |  | - |
| **Lung function**<br>**Mean (sd)** |  |  |  |  |
| **FEV1** | 81.39 (18.94) | 91.35 (14.85) | 1.26e-18 |  |
| **FEV1/FVC** | 72.72 (8.29) | 77.79 (7.88) | 2.05e-02 |  |
| **FEV1/VC** | 71.62 (12.78) | 75.59 (12.96) | 4.37e-06 |  |
| **FVC** | 86.50 (17.26) | 94.67 (14.12) | 4.93e-13 |  |
| **VC** | 86.08 (16.50) | 93.43 (14.6) | 2.69e-12 |  |
| **DLCO** | 74.77 (27.42) | 83.74 (23.34) | 1.07e-10 |  |
| **TLC** | 83.39 (23.76) | 89.82 (26.27) | 4.87e-12 |  |
| **Löfgren's syndrome (LS) (n = 487)** | 77 | 410 |  |  |
| % males | 67.5% | 57.1% | 0.101 |  |
| % females | 32.5% | 42.9% |  |  |
| **Age (years)** (Mean ± SD) | 44.14 ± 12.7 | 40.14 ± 11.09 | 0.0049 |  |
| **Lung function in LS**<br>**Median (sd)** |  |  |  |  |
| **FEV1** | 84.5 (15) | 92 (14.09) | 0.007 |  |
| **FEV1/FVC** | 78.5 (7.14) | 78.5 (7.70) | 0.850 |  |
| **FEV1/VC** | 77 (7.74) | 77 (13.49) | 0.921 |  |
| **FVC** | 89 (15) | 96 (13.63) | 0.015 |  |
| **VC** | 89 (13.65) | 95 (14.49) | 0.006 |  |
| **DLCO** | 85 (21.76) | 88 (22.6) | 0.526 |  |
| **TLC** | 93 (10.59) | 96 (26.15) | 0.114 |  |
| **Non-Löfgren's sarcoidosis (non-LS) (n = 997)** | 698 | 299 |  |  |
| % males | 65.8% | 54.5% | 1.01e-03 |  |
| % females | 34.2% | 45.5% |  |  |
| **Age (years)** (Mean ± SD) | 45.62 ± 11.61 | 43.75 ± 12.54 |  |  |
| **Lung function in nonLS**<br>**Median (IQR)** |  |  |  |  |
| **FEV1 %** | 81 (19.2) | 90 (15.89) | 1.98e-07 |  |
| **FEV1/FVC %** | 73 (8.34) | 75.5 (8.65) | 2.65e-01 |  |
| **FEV1/VC %** | 73 (13.10) | 76 (12.07) | 2.68e-03 |  |
| **FVC %** | 86 (17.44) | 95 814.94) | 8.72e-06 |  |
| **VC %** | 86 (16.76) | 92 (14.60 | 1.44e04 |  |
| **DLCO** | 78 (27.75) | 85 (24.46) | 2.56e-04 |  |
| **TLC** | 87 (24.55) | 92.5 (26.55) | 3.42e-05 |  |

**Table 2.** Results from logistic regression on chronic versus nonchronic.

| ***HLA-DRB1* allele** | **OR** | **95% CI lower** | **95% CI upper** | **P-value** | **FDR** |
|---|---|---|---|---|---|
| *\*01* | 0.545 | 0.390 | 0.756 | 3.17E-04 | **1.03E-03** |
| *\*03* | 0.147 | 0.113 | 0.191 | 5.74E-46 | **7.46E-45** |
| *\*04* | 1.160 | 0.908 | 1.482 | 2.36E-01 | 3.40E-01 |
| *\*07* | 2.130 | 1.465 | 3.137 | 9.50E-05 | **4.12E-04** |
| *\*08* | 1.787 | 1.213 | 2.665 | 3.72E-03 | **6.91E-03** |
| *\*09* | 1.368 | 0.743 | 2.571 | 3.18E-01 | 4.14E-01 |
| *\*10* | 1.155 | 0.513 | 2.650 | 7.27E-01 | 7.27E-01 |
| *\*11* | 1.669 | 1.191 | 2.357 | 3.22E-03 | **6.91E-03** |
| *\*12* | 2.031 | 1.252 | 3.375 | 4.90E-03 | **7.96E-03** |
| *\*13* | 0.916 | 0.712 | 1.177 | 4.92E-01 | 5.82E-01 |
| *\*14* | 2.060 | 1.345 | 3.211 | 1.09E-03 | **2.83E-03** |
| *\*15* | 1.945 | 1.533 | 2.472 | 4.78E-08 | **3.11E-07** |
| *\*16* | 0.729 | 0.239 | 2.107 | 5.60E-01 | 6.06E-01 |

The median age in the chronic group was 45 (±11.4), and in the nonchronic group, 39 (±11.6). Biological sex distribution differed between chronic and nonchronic groups (X-squared = 14.99, df = 1, p = 1.08e-4). The chronic group had 56.3% males and 45.8% females, and the nonchronic group had 43.7% males and 54.2% females. Moreover, regression analysis by sex showed that males had higher odds (OR = 1.52; 95% CI: 1.23-1.87, P=8.96e-05) of chronic disease than females. In the healthy control group (n = 5,817), the median age at recruitment was 55 (±11.7) years, with 27.5% male.

Further characterization of the disease trajectory by clinical phenotype showed notable differences between chronic and non-chronic cases within the LS and non-LS groups. Regression analysis stratified by sex revealed that males in the LS group (OR = 1.56, 95% CI: 0.94 - 2.61, P=8.88e-2) and males in the non-LS group (OR = 1.60, 95% CI: 1.21 - 2.11, P=8.18e-4) exhibited significantly increased odds of having a chronic disease compared to their female counterparts, respectively.

### *HLA-DRB1* analysis

Figure 2 illustrates the distribution of *HLA-DRB1* alleles among chronic and nonchronic phenotypes. The Chi-square test identified *HLA-DRB1 *01, *03, *07, *08, *11, *12, *14*, and *\*15* as the most relevant alleles after applying multiple-comparison correction (FDR < 0.05).

**Figure 2.**
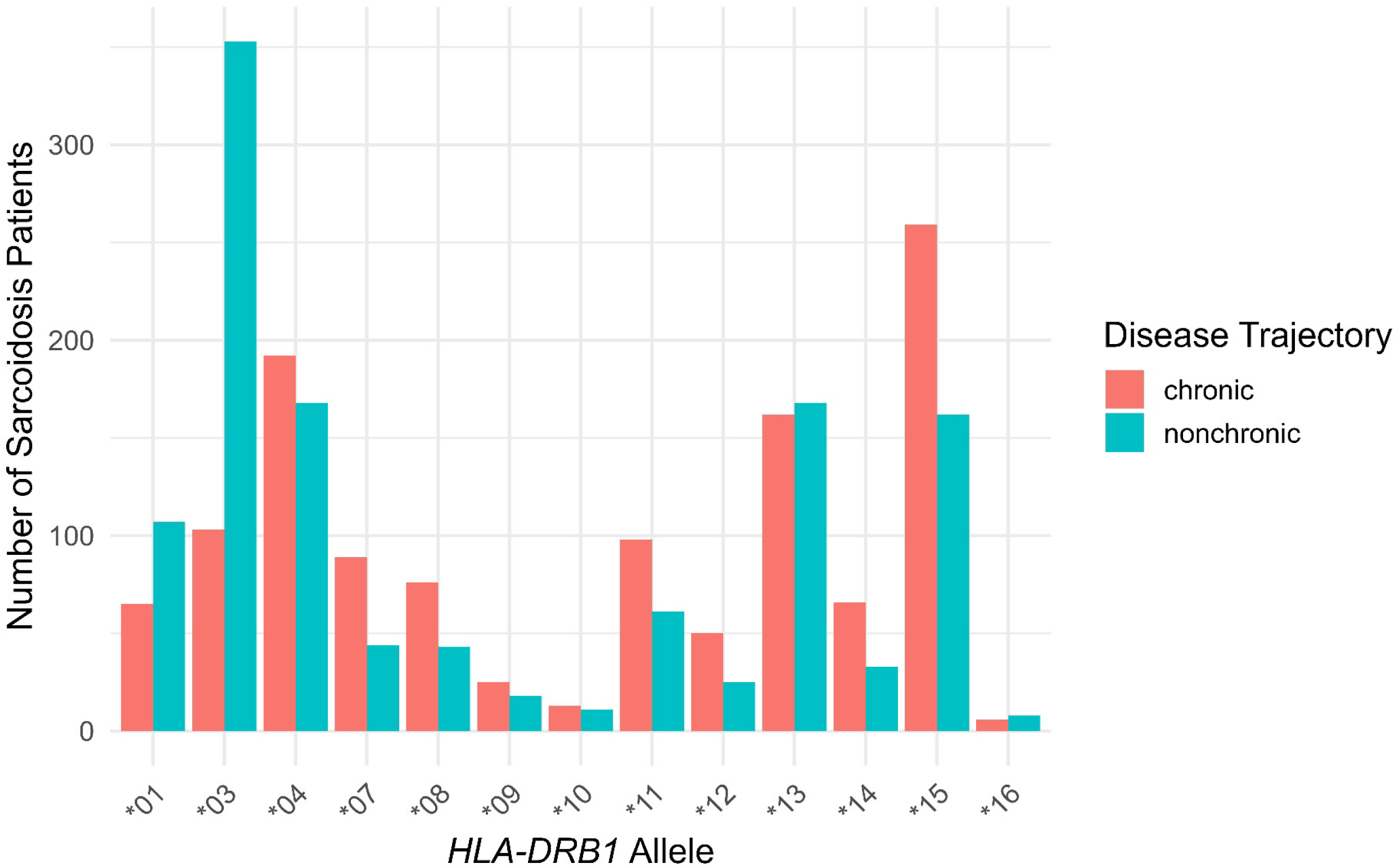
HLA-DRB1 distribution by disease trajectory

Among clinical phenotypes LS and nonLS, the relevance of *HLA-DRB1* alleles varied considerably between disease trajectory groups. Specifically, in LS, the distribution of alleles *\*03, *04,* and *\*15* was most relevant when comparing chronic versus nonchronic groups, while in nonLS, the distribution of alleles *\*03* and *\*07* was the most relevant in disease trajectory comparisons. Details of the distribution of *HLA-DRB1* alleles can be found in Supplementary Table S1 A-C.

Logistic regression analysis of the impact of *HLA-DRB1* alleles on disease trajectory showed lower odds of chronic disease than nonchronic disease for patients carrying *HLA-DRB1*01* (OR = 0.545, 95% CI: 0.390-0.756) and *\*03* (OR = 0.147, 95% CI: 0.133-0.191). Conversely, *HLA-DRB1* alleles *\*07, *08, *11, *12, *14,* and *\*15* were associated with higher odds of chronic disease compared with nonchronic disease (Table 1).

Notably, *HLA-DRB1*03* was associated with lower odds of chronic disease, regardless of LS or nonLS diagnosis. This pattern was not evident for the other alleles across clinical groups (Supplementary Table S1 D).

### Association analyses

We observed genetic differences in disease trajectory phenotypes, chronic and nonchronic. In the chronic phenotype, we identified three genomic regions marked by three lead signals and 12 independent genetic variants. The lead signals were rs3135387 (OR = 1.659, 95% CI 1.436–1.916) near *HLA-DRA*, rs3117224 (OR = 1.623, 95% CI 1.396–1.887) located 2.2 kb 3′ of *HLA-DPB1*, and rs3762097 (OR = 0.622, 95% CI 0.538–0.718) in an intron of *PLAC9*. In the nonchronic phenotype, one genomic locus was predominant, with 35 lead signals and 370 independent genetic variants. The lead signal was rs2849018 (OR = 3.70, 95% CI 3.14–4.37) in an intron of *TSBP1* on chromosome 6 (Table 3). Complete significant results for chronic and nonchronic sarcoidosis are in Supplementary Tables S2-A and S2-B, respectively. Figure 4 shows the corresponding Manhattan plots.

**Table 3.** GWAS findings for chronic and nonchronic disease trajectory phenotypes.

| Phenotype | Genomic Locus marked by uniqID | rsID | chr | Pos (hg19) | P-value (GC) | start | end | N SNPs | N GWAS SNPs | N Ind SigSNPs | N Lead SNPs |
|---|---|---|---|---|---|---|---|---|---|---|---|
| **Chronic** |  |  |  |  |  |  |  |  |  |  |  |
|  | 6:32415109:G:T | rs3135387 | 6 | 32415109 | 2.21E-11 | 32340106 | 32632832 | 652 | 247 | 5 | 1 |
|  | 6:33059625:C:G | rs3117224 | 6 | 33059625 | 8.91E-10 | 33041064 | 33194477 | 385 | 208 | 3 | 1 |
|  | 10:81901375:A:C | rs3762097 | 10 | 81901375 | 3.05E-10 | 81895839 | 82210206 | 203 | 155 | 4 | 1 |
| **nonchronic** |  |  |  |  |  |  |  |  |  |  |  |
|  | 6:32310863:C:T | rs2849018 | 6 | 32310863 | 1.835e-50 | 25182925 | 33196570 | 13430 | 7297 | 370 | 35 |

uniqID = unique genomic-locus identifier; rsID = reference SNP identifier; hg19 = human genome assembly 19; P value (GC) = genomic-control-adjusted P value; N SNPs = number of SNPs within the genomic locus; N GWAS SNPs = number of SNPs meeting the analysis-specific FUMA threshold; N Ind Sig SNPs = number of independent significant SNPs; N Lead SNPs = number of lead SNPs.

Among LS and non-LS sarcoidosis, we identified additional genetic variants associated with the chronic and nonchronic phenotypes. In LS nonchronic sarcoidosis, we identified one genome-wide significant locus led by rs2849018 (OR = 5.90, 95% CI 4.83–7.22) within an intron of *TSBP1*, with 47 lead signals and 530 independent variants (Table 4). In LS chronic sarcoidosis, 11 suggestive loci met the exploratory FUMA threshold of P < 5 × 10⁻⁶: rs78180666 (OR = 3.54, 95% CI 2.09–6.00), rs6837980 (OR = 3.65, 95% CI 2.14–6.22), rs118158252 (OR = 3.36, 95% CI 2.02–5.60), rs73188811 (OR = 3.58, 95% CI 2.13–6.01), rs17474411 (OR = 3.29, 95% CI 2.01–5.42), rs1227084 (OR = 3.55, 95% CI 2.11–5.99), rs71488732 (OR = 2.65, 95% CI 1.75–4.02), rs17099622 (OR = 3.94, 95% CI 2.37–6.55), rs4408393 (OR = 2.87, 95% CI 1.94–4.24), rs113886122 (OR = 2.90, 95% CI 1.89–4.44), and rs4488763 (OR = 2.918, 95% CI 1.853–4.596). However, none reached P < 5 × 10⁻⁸; the minimum genomic-control-adjusted P value was 1.226 × 10⁻⁷. Supplementary Table S3 shows complete significant results for disease trajectory stratified by LS.

**Table 4.** GWAS association results for LS and non-LS by disease trajectory phenotypes.

| **Sarcoidosis clinical phenotypes** | **Genomic Locus marked by uniqID** | **rsID** | **chr** | **Pos (hg19)** | **P-value (GC)** | **start** | **end** | **N SNPs** | **N GWAS SNPs** | **N Ind SigSNPs** | **N Lead SNPs** |
|---|---|---|---|---|---|---|---|---|---|---|---|
| **LS chronic** |  |  |  |  |  |  |  |  |  |  |  |
| **1** | 3:195895972:C:T | rs78180666 | 3 | 195895972 | 2.52E-06 | 195895972 | 195895972 | 1 | 1 | 1 | 1 |
| **2** | 4:24213123:C:T | rs6837980 | 4 | 24213123 | 2.05E-06 | 24141861 | 24220057 | 31 | 10 | 1 | 1 |
| **3** | 6:153693559:A:G | rs118158252 | 6 | 153693559 | 3.34E-06 | 153664453 | 153694723 | 8 | 5 | 1 | 1 |
| **4** | 8:5024826:C:T | rs73188811 | 8 | 5024826 | 1.42E-06 | 5024826 | 5024826 | 1 | 1 | 1 | 1 |
| **5** | 8:15591606:A:C | rs17474411 | 8 | 15591606 | 2.45E-06 | 15551828 | 15591606 | 2 | 1 | 1 | 1 |
| **6** | 10:73449596:A:G | rs1227084 | 10 | 73449596 | 2.01E-06 | 73442952 | 73449596 | 6 | 3 | 1 | 1 |
| **7** | 11:20158097:C:T | rs71488732 | 11 | 20158097 | 4.80E-06 | 20155352 | 20159603 | 7 | 6 | 1 | 1 |
| **8** | 11:102700352:A:G | rs17099622 | 11 | 102700352 | 1.33E-07 | 102700352 | 102727831 | 3 | 3 | 1 | 1 |
| **9** | 12:103797108:C:T | rs4408393 | 12 | 103797108 | 1.23E-07 | 103775694 | 103895792 | 150 | 113 | 2 | 1 |
| **10** | 19:50044741:C:G | rs113886122 | 19 | 50044741 | 9.64E-07 | 50044741 | 50051561 | 2 | 1 | 1 | 1 |
| **11** | 22:32380164:C:G | rs4488763 | 22 | 32380164 | 3.83E-06 | 32380164 | 32380164 | 1 | 1 | 1 | 1 |
| **LS nonchronic** |  |  |  |  |  |  |  |  |  |  |  |
| **1** | 6:32310863:C:T | rs2849018 | 6 | 32310863 | 6.28E-65 | 25182925 | 33196570 | 17300 | 9495 | 530 | 47 |
| **non-LS chronic** |  |  |  |  |  |  |  |  |  |  |  |
| **1** | 6:32415109:G:T | rs3135387 | 6 | 32415109 | 2.96E-10 | 32340106 | 32632832 | 417 | 222 | 4 | 1 |
| **2** | 6:33059625:C:G | rs3117224 | 6 | 33059625 | 2.69E-10 | 33041064 | 33194477 | 385 | 208 | 3 | 1 |
| **3** | 10:81910278:A:G | rs7071579 | 10 | 81910278 | 1.08E-08 | 81899632 | 82210206 | 121 | 89 | 2 | 1 |
| **non-LS nonchronic** |  |  |  |  |  |  |  |  |  |  |  |
| **1** | 3:12908760:C:T | rs76164113 | 3 | 12908760 | 5.20E-07 | 12908760 | 12910538 | 2 | 1 | 1 | 1 |
| **2** | 6:87877896:A:G | rs59344869 | 6 | 87877896 | 3.57E-07 | 87815232 | 88013881 | 115 | 1 | 1 | 1 |
| **3** | 6:99750744:A:T | rs57019067 | 6 | 99750744 | 2.60E-06 | 99700069 | 99871830 | 23 | 19 | 1 | 1 |
| **4** | 6:123605648:A:G | rs4273711 | 6 | 123605648 | 1.41E-06 | 123571228 | 123669127 | 7 | 4 | 1 | 1 |
| **5** | 8:3658428:A:T | rs112185026 | 8 | 3658428 | 1.90E-06 | 3620205 | 3670511 | 54 | 52 | 1 | 1 |
| **6** | 14:42189017:A:T | rs12432259 | 14 | 42189017 | 7.21E-07 | 41965619 | 42203673 | 3 | 2 | 1 | 1 |
| **7** | 21:32295397:C:T | rs1557278 | 21 | 32295397 | 8.62E-07 | 32291501 | 32303565 | 12 | 8 | 1 | 1 |

uniqID = unique genomic-locus identifier; rsID = reference SNP identifier; hg19 = human genome assembly 19; P value (GC) = genomic-control-adjusted P value; N SNPs = number of SNPs within the genomic locus; N GWAS SNPs = number of SNPs meeting the analysis-specific FUMA threshold; N Ind Sig SNPs = number of independent significant SNPs; N Lead SNPs = number of lead SNPs.

In non-LS chronic sarcoidosis, three genome-wide significant loci were led by rs3135387 (OR = 1.649, 95% CI 1.418–1.918), rs3117224 (OR = 1.683, 95% CI 1.439–1.968), and rs7071579 (OR = 1.539, 95% CI 1.333–1.776). In non-LS nonchronic sarcoidosis, seven suggestive loci met the exploratory FUMA threshold of P < 5 × 10⁻⁶: rs76164113 (OR = 2.442, 95% CI 1.738–3.432), rs59344869 (OR = 2.424, 95% CI 1.738–3.380), rs57019067 (OR = 2.162, 95% CI 1.580–2.960), rs4273711 (OR = 1.724, 95% CI 1.389–2.139), rs112185026 (OR = 2.188, 95% CI 1.598–2.995), rs12432259 (OR = 2.591, 95% CI 1.794–3.740), and rs1557278 (OR = 1.839, 95% CI 1.451–2.330), but none reached P < 5 × 10⁻⁸; the minimum genomic-control-adjusted P value was 3.572 × 10⁻⁷. Supplementary Table S4 shows complete significant results for disease trajectory stratified by nonLS. Supplementary Figures S2 and S3 show Manhattan plots for chronic and nonchronic cases stratified by LS and non-LS, respectively.

### SNP Heritability analyses

SNP heritability for disease trajectory phenotypes was estimated at 32.8% (95% CI 23.9–41.7%) for chronic sarcoidosis and 47.5% (95% CI 37.5–57.6%) for nonchronic sarcoidosis (Figure 3). Within the clinical subgroups, SNP heritability was 37.2% (95% CI 27.1–47.2%) for non-LS chronic, 57.1% (95% CI 45.2–68.9%) for LS nonchronic, and 71.7% (95% CI 46.4–97.0%) for non-LS nonchronic sarcoidosis. The LS chronic estimate was 20.7% (95% CI −35.2–76.7%); however, its reliability is limited by the small sample size in this group (Supplementary Figure S1).

**Figure 3.**
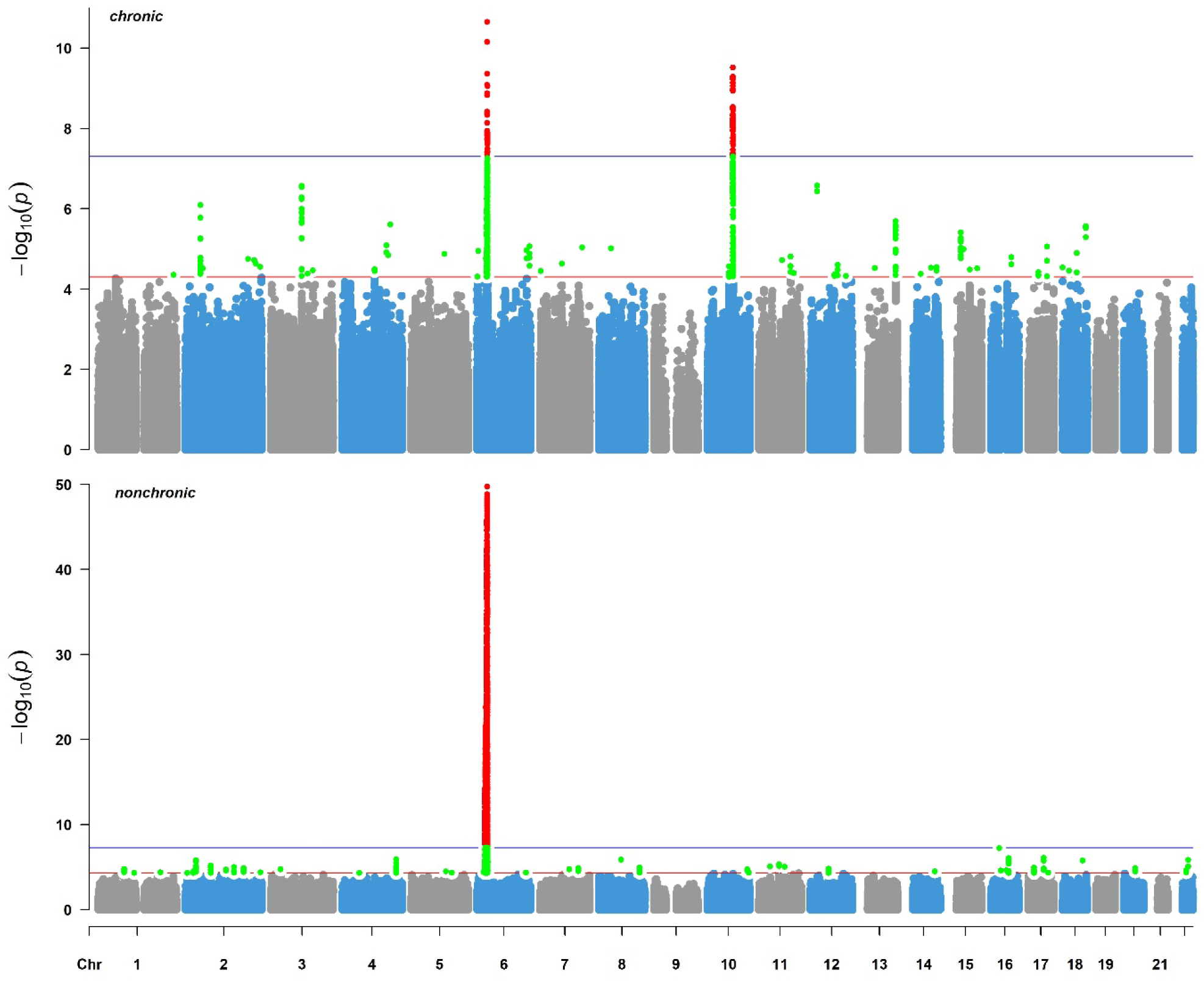
Manhattan plots for chronic (top) and nonchronic (bottom) disease trajectory phenotypes

**Figure 4.**
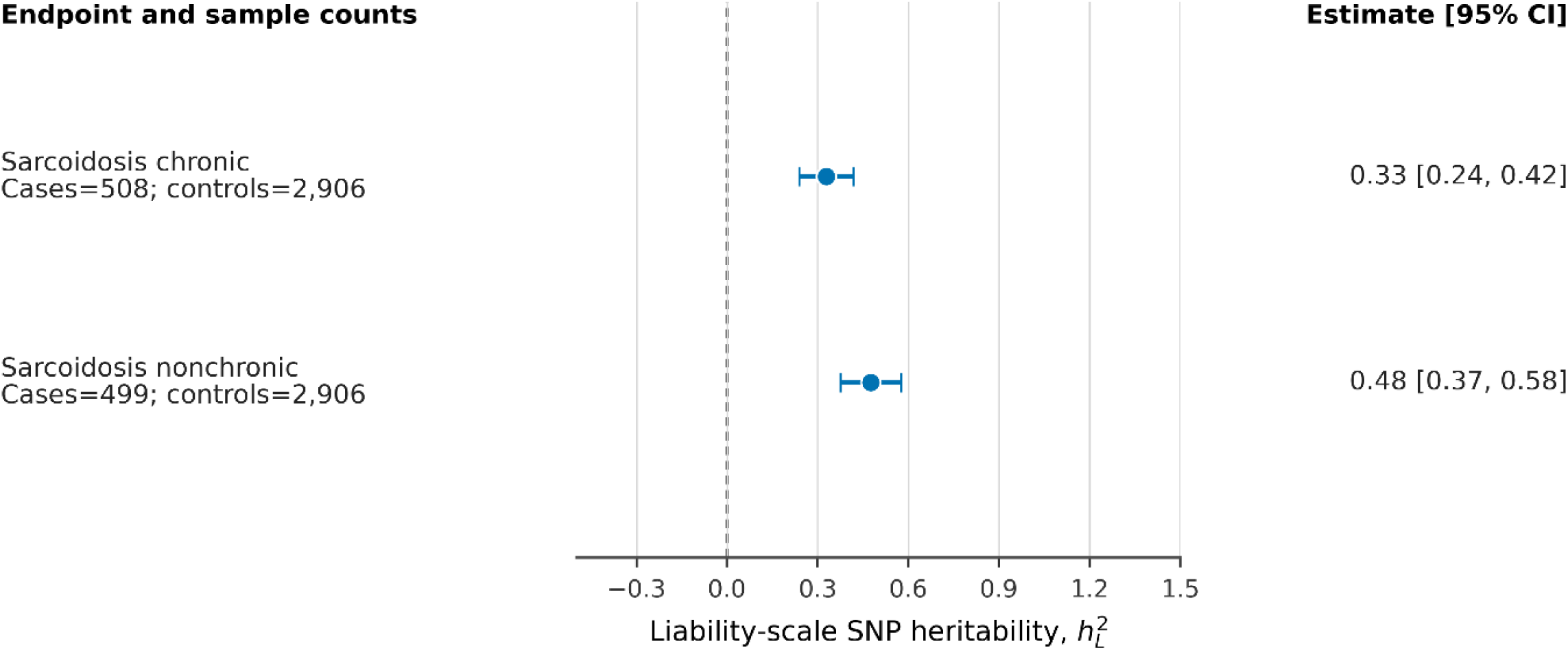
Forest plot showing SNP heritability in chronic and nonchronic sarcoidosis

### Gene-based analyses

Gene-based analyses identified 12 genes associated with the chronic phenotype (*TSBP1, BTNL2, HLA-DRA, HLA-DRB1, HLA-DQA1, HLA-DPA1, RING1, RXRB* on chromosome 6 and *PLAC9, ANXA11, DYDC2, TSPAN14* on chromosome 10) (Table 5) at P < 2.687 × 10⁻⁶. Likewise, we identified 205 genes associated with the nonchronic phenotype, mostly in the extended MHC region on chromosome 6 (25–33 Mb) (Table 6, Supplementary Table S5). Gene-based analyses stratified by Löfgren’s syndrome identified 223 significant genes in LS nonchronic sarcoidosis, chiefly 222 on chromosome 6 and *PTK2B* on chromosome 8 (Supplementary Table S6 A). LS chronic sarcoidosis had one significant gene, *C12orf42* on chromosome 12 (Supplementary Table S6 B). Non-LS chronic sarcoidosis had 11 significant genes, chiefly nine on chromosome 6 and two genes, *ANXA11* and *TSPAN14,* on chromosome 10 (Supplementary Table S7 A). In non-LS nonchronic sarcoidosis, *GMEB1* was the most prominent gene (P = 1.4655 × 10⁻⁵); however, following Bonferroni correction, no genes remained statistically significant (Supplementary Table S7 B).

**Table 5.** Results from gene-based analyses for chronic phenotype.

| GENE | CHR | START | STOP | NSNPS | NPARAM | N | ZSTAT | P | SYMBOL |
|---|---|---|---|---|---|---|---|---|---|
| **ENSG00000204296** | 6 | 32246303 | 32374684 | 823 | 20 | 6316 | 5.2261 | **8.66E-08** | *TSBP1* |
| **ENSG00000204290** | 6 | 32351740 | 32409905 | 519 | 18 | 6320 | 5.9633 | **1.24E-09** | *BTNL2* |
| **ENSG00000204287** | 6 | 32372619 | 32422823 | 494 | 21 | 6320 | 5.5759 | **1.23E-08** | *HLA-DRA* |
| **ENSG00000196126** | 6 | 32536546 | 32592625 | 122 | 10 | 6311 | 4.8628 | **5.79E-07** | *HLA-DRB1* |
| **ENSG00000196735** | 6 | 32560956 | 32624839 | 175 | 12 | 6296 | 4.6823 | **1.42E-06** | *HLA-DQA1* |
| **ENSG00000231389** | 6 | 33022346 | 33083552 | 550 | 10 | 6304 | 5.1287 | **1.46E-07** | *HLA-DPA1* |
| **ENSG00000204227** | 6 | 33141272 | 33190499 | 80 | 11 | 6308 | 4.8737 | **5.48E-07** | *RING1* |
| **ENSG00000204231** | 6 | 33151365 | 33203630 | 75 | 12 | 6280 | 5.1604 | **1.23E-07** | *RXRB* |
| **ENSG00000189129** | 10 | 81856438 | 81915115 | 191 | 24 | 6179 | 4.9736 | **3.29E-07** | *PLAC9* |
| **ENSG00000122359** | 10 | 81900645 | 82000328 | 254 | 16 | 6275 | 5.3678 | **3.99E-08** | *ANXA11* |
| **ENSG00000204038** | 10 | 81974466 | 82023395 | 102 | 11 | 6277 | 5.1554 | **1.27E-07** | *DYDC2* |
| **ENSG00000108219** | 10 | 82178922 | 82302879 | 224 | 18 | 6254 | 4.9437 | **3.83E-07** | *TSPAN14* |

**Table 6.** Results from gene-based analyses for nonchronic phenotype (complete results are shown in Supplementary Table 5)

| GENE | CHR | START | STOP | NSNPS | NPARAM | N | ZSTAT | P | SYMBOL |
|---|---|---|---|---|---|---|---|---|---|
| **ENSG00000124568** | 6 | 25773125 | 25867287 | 211 | 14 | 6299 | 5.5034 | **1.86E-08** | *SLC17A1* |
| **ENSG00000198366** | 6 | 25985718 | 26031186 | 71 | 9 | 6291 | 5.5135 | **1.76E-08** | *HIST1H3A* |
| **ENSG00000181315** | 6 | 26626518 | 26694980 | 100 | 8 | 6305 | 5.4398 | **2.67E-08** | *ZNF322* |
| **ENSG00000168131** | 6 | 27868963 | 27915174 | 35 | 6 | 6300 | 6.5379 | **3.12E-11** | *OR2B2* |
| **ENSG00000196812** | 6 | 28057338 | 28107860 | 122 | 7 | 6308 | 7.2971 | **1.47E-13** | *ZSCAN16* |
| **ENSG00000204713** | 6 | 28860779 | 28926766 | 148 | 10 | 6291 | 6.2564 | **1.97E-10** | *TRIM27* |
| **ENSG00000137312** | 6 | 30685486 | 30745510 | 102 | 13 | 6302 | 6.1094 | **5.00E-10** | *FLOT1* |
| **ENSG00000204544** | 6 | 30916495 | 30967680 | 175 | 13 | 6296 | 5.7141 | **5.51E-09** | *MUC21* |
| **ENSG00000213719** | 6 | 31688358 | 31742540 | 66 | 10 | 6307 | 6.1094 | **5.00E-10** | *CLIC1* |
| **ENSG00000204385** | 6 | 31820969 | 31881823 | 85 | 11 | 6303 | 7.2792 | **1.68E-13** | *SLC44A4* |
| **ENSG00000166278** | 6 | 31830562 | 31923449 | 126 | 17 | 6305 | 6.8429 | **3.88E-12** | *C2* |
| **ENSG00000231852** | 6 | 31971042 | 32019447 | 14 | 5 | 6200 | 6.7457 | **7.62E-12** | *CYP21A2* |
| **ENSG00000168477** | 6 | 31998931 | 32118111 | 135 | 21 | 6296 | 6.1094 | **5.00E-10** | *TNXB* |
| **ENSG00000213676** | 6 | 32055953 | 32131030 | 91 | 18 | 6301 | 7.8242 | **2.55E-15** | *ATF6B* |
| **ENSG00000221988** | 6 | 32086218 | 32144011 | 76 | 14 | 6303 | 6.1094 | **5.00E-10** | *PPT2* |
| **ENSG00000204310** | 6 | 32125989 | 32180873 | 110 | 15 | 6304 | 6.1094 | **5.00E-10** | *AGPAT1* |
| **ENSG00000204305** | 6 | 32138745 | 32187101 | 114 | 16 | 6304 | 6.1094 | **5.00E-10** | *AGER* |
| **ENSG00000204304** | 6 | 32142512 | 32192963 | 141 | 19 | 6304 | 6.1094 | **5.00E-10** | *PBX2* |
| **ENSG00000204301** | 6 | 32152620 | 32226844 | 354 | 31 | 6305 | 8.1738 | **1.49E-16** | *NOTCH4* |
| **ENSG00000204296** | 6 | 32246303 | 32374684 | 823 | 20 | 6307 | 8.2345 | **9.01E-17** | *TSBP1* |
| **ENSG00000204290** | 6 | 32351740 | 32409905 | 519 | 18 | 6311 | 7.5772 | **1.77E-14** | *BTNL2* |
| **ENSG00000204287** | 6 | 32372619 | 32422823 | 494 | 21 | 6311 | 6.6906 | **1.11E-11** | *HLA-DRA* |
| **ENSG00000196126** | 6 | 32536546 | 32592625 | 122 | 10 | 6301 | 6.1094 | **5.00E-10** | *HLA-DRB1* |
| **ENSG00000196735** | 6 | 32560956 | 32624839 | 175 | 12 | 6287 | 7.6716 | **8.49E-15** | *HLA-DQA1* |
| **ENSG00000179344** | 6 | 32617244 | 32671160 | 282 | 14 | 6303 | 7.2811 | **1.66E-13** | *HLA-DQB1* |
| **ENSG00000237541** | 6 | 32674119 | 32724992 | 605 | 20 | 6303 | 7.9179 | **1.21E-15** | *HLA-DQA2* |
| **ENSG00000232629** | 6 | 32713875 | 32766311 | 529 | 14 | 6299 | 7.1327 | **4.92E-13** | *HLA-DQB2* |
| **ENSG00000250264** | 6 | 32771544 | 32841599 | 381 | 31 | 6311 | 8.6983 | **1.69E-18** | *TAP2* |
| **ENSG00000240065** | 6 | 32776913 | 32837362 | 298 | 30 | 6310 | 8.122 | **2.29E-16** | *PSMB9* |
| **ENSG00000223865** | 6 | 33008703 | 33064978 | 509 | 14 | 6291 | 6.5887 | **2.22E-11** | *HLA-DPB1* |

### Functional mapping and annotations

eQTL enrichment using FUMA and multiple eQTL datasets revealed several eQTL SNPs. In particular, we identified several eQTL SNPs as regulatory markers of gene expression associated with GWAS signals, particularly in lung tissue, whole blood, and immune cell populations. eQTL SNPs in monocytes, B cells, and neutrophils were more prevalent in the chronic phenotype (Figure 6), whereas a higher immune cell diversity profile, including monocytes, macrophages, T cells, and B cells, was present in the nonchronic phenotype (Figure 7). We also detected eQTL SNPs in various other organs and tissues, shown in Supplementary Figures S8 A and S8 B for chronic and nonchronic, respectively.

**Figure 5.**
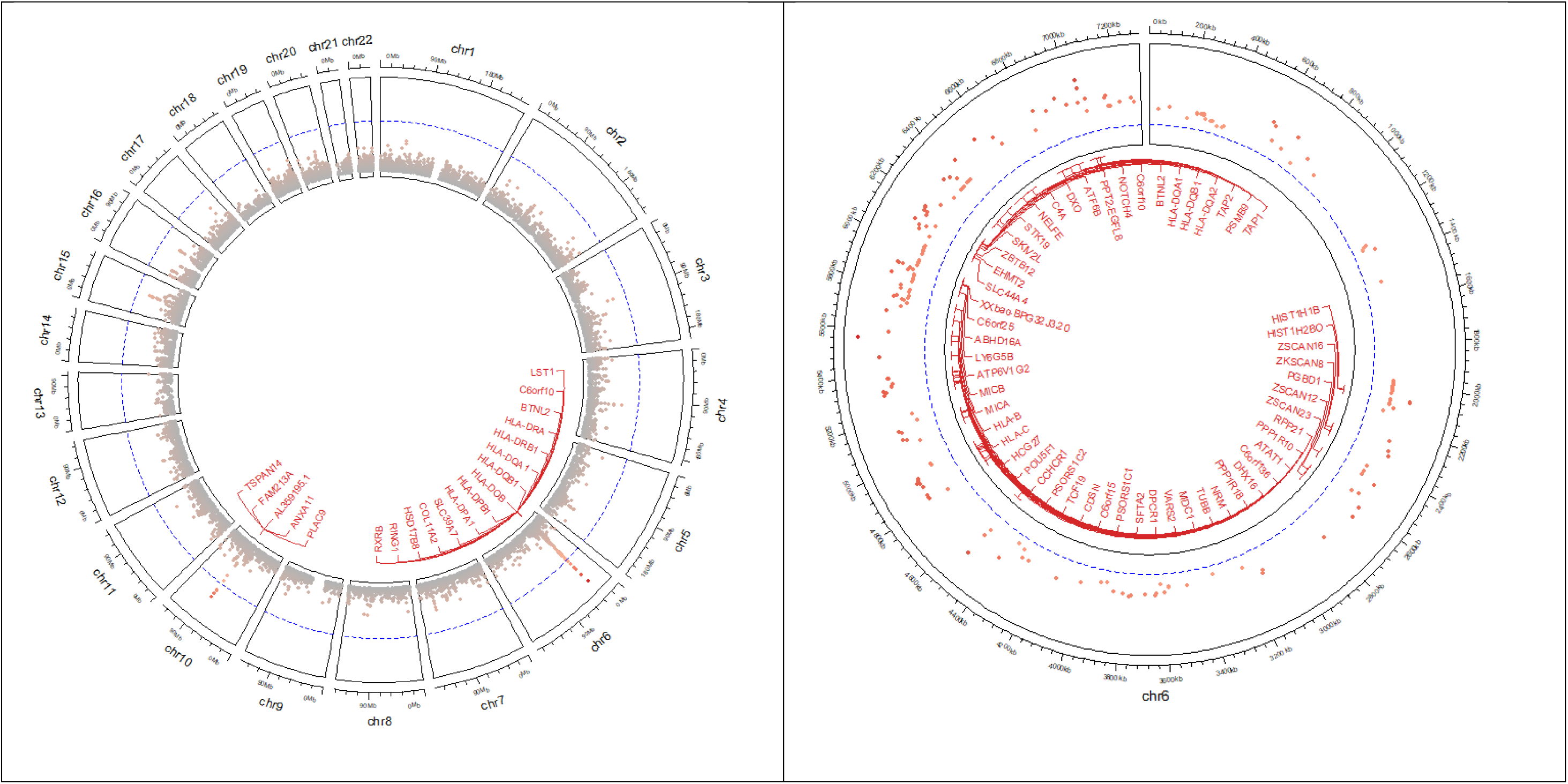
Mapping results from gene-based analyses for chronic (left) and nonchronic (right) phenotypes *Note: The gene-based analyses for nonchronic include 205 genes, but the Circos plot displays only the top 50 genes. Full results are available in Supplementary Table S5*.

**Figure 6.**
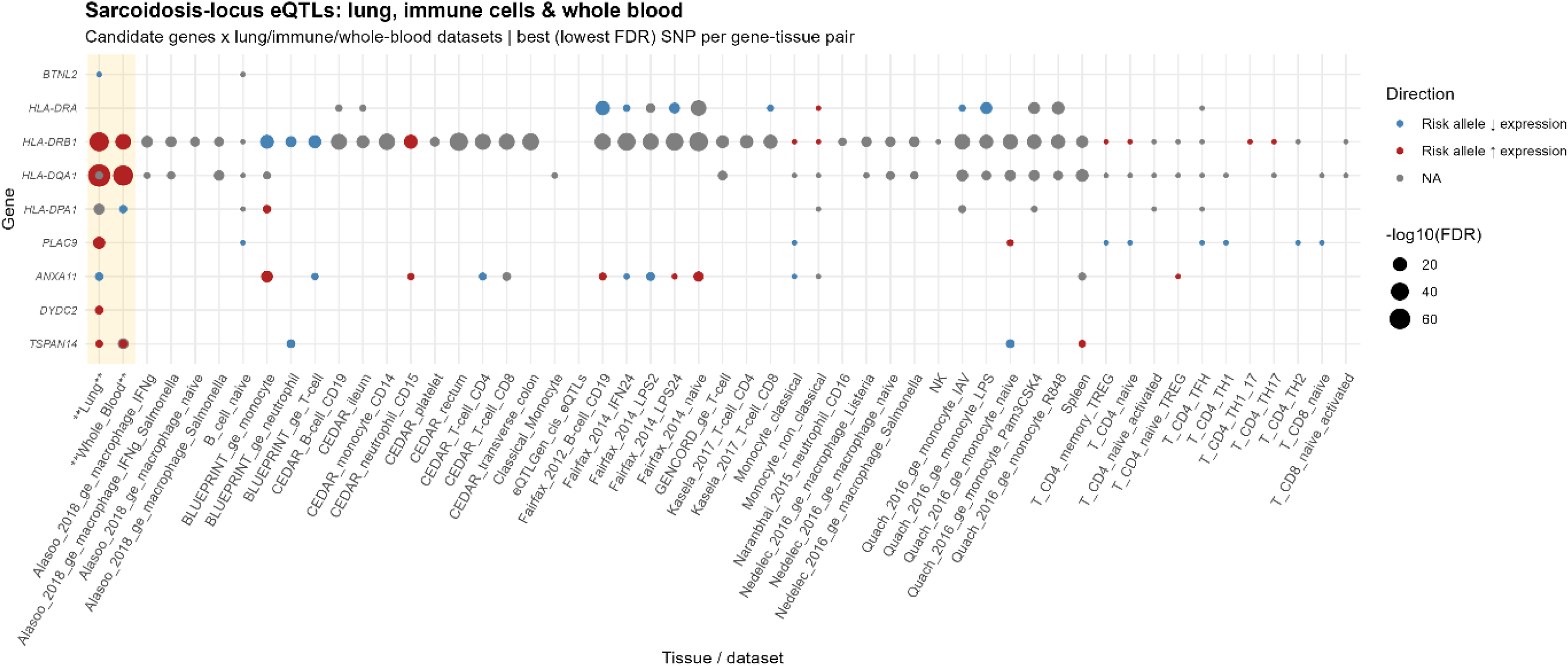
eQTL mapping in emphasized tissues and cell types for chronic sarcoidosis

**Figure 7.**
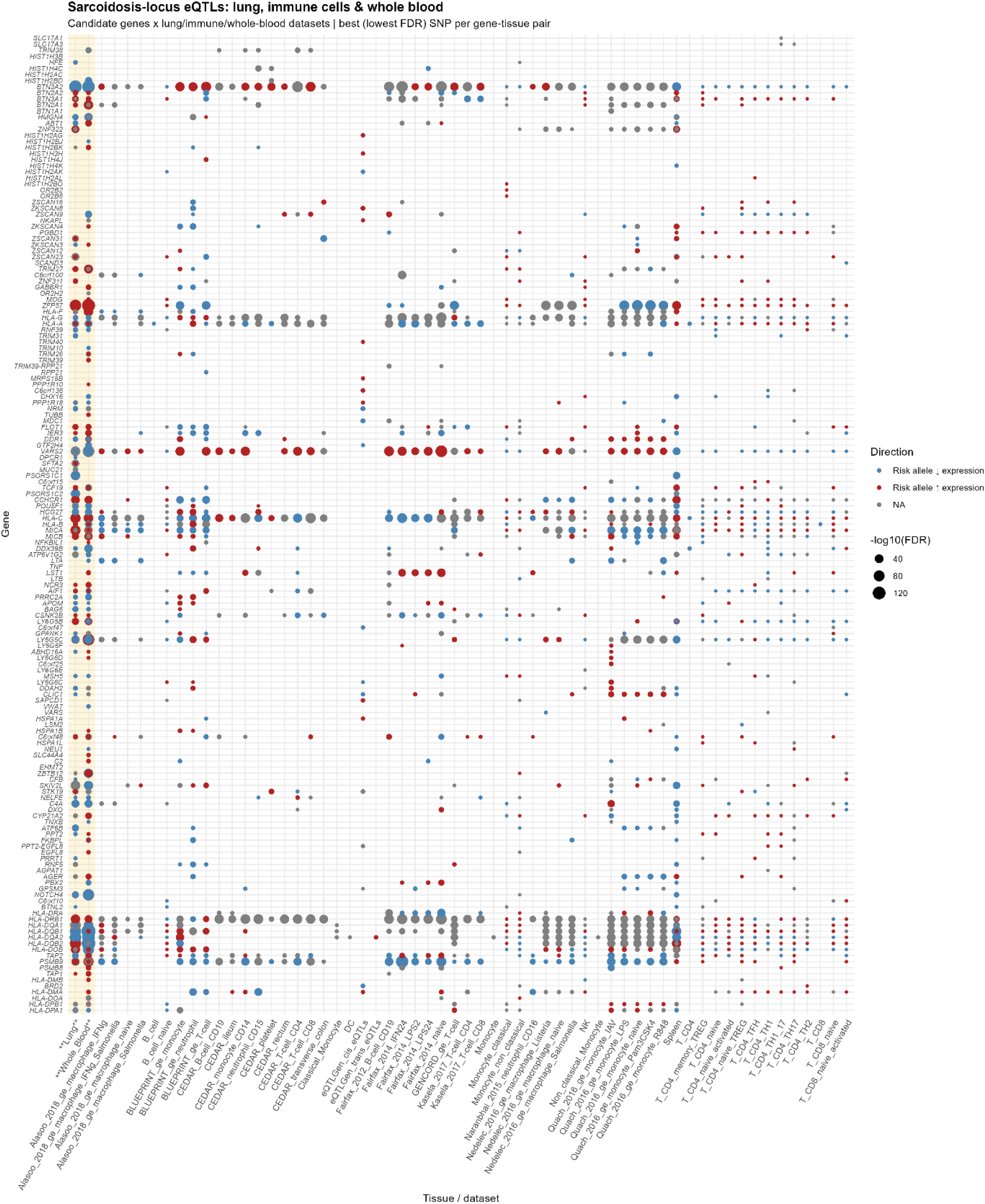
eQTL mapping in emphasized tissues and cell types for nonchronic sarcoidosis

### Pathway analyses

IPA identified the antigen presentation pathway as the leading signaling pathway for chronic sarcoidosis (16 genes; P = 2.00 × 10^−11^) (Supplementary Table S9 A) and nonchronic sarcoidosis (23 genes; P = 7.94 × 10^−20^) (Supplementary Table S9 B). Given the importance of immune-mediated genes, we identified over 100 pathways, some specific to each disease trajectory phenotype and others shared between them.

Across clinical phenotypes, we also identified different signaling pathways. In LS nonchronic, the antigen presentation pathway (23 genes; P = 2.00 × 10^−20^) was the most significant (Supplementary Table S10 A). In LS chronic, IL-17A signaling in airway cells (10 genes; P = 3.31 × 10^−4^) was the top nominal pathway (Supplementary Table S10 B). In non-LS chronic sarcoidosis, the antigen presentation pathway (15 genes; P = 1.95 × 10⁻¹⁰) was a top finding (Supplementary Table S11 A). In non-LS nonchronic sarcoidosis, the interferon gamma signaling pathway (13 genes; P = 5.75 × 10^−4^) was the top nominal pathway (Supplementary Table S11 B). Figure 8 shows the top fifteen pathways across chronic and nonchronic subtypes.

**Figure 8.**
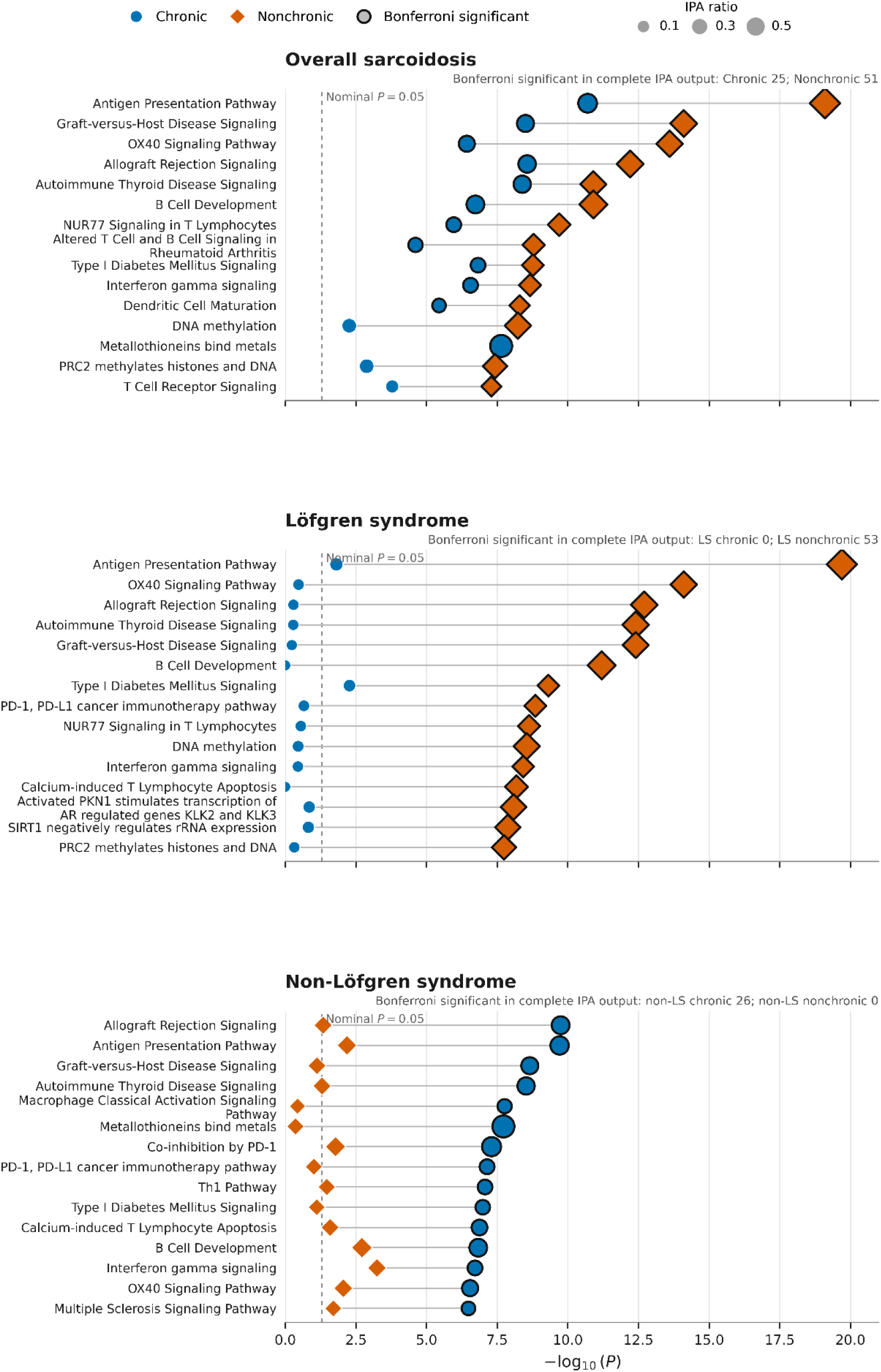
IPA canonical pathway results across sarcoidosis trajectory phenotypes

## Discussion

We analyzed the genetics of disease trajectory in sarcoidosis, focusing on both chronic and nonchronic phenotypes. Our study revealed differences in the genetic architecture between these groups. Moreover, by stratifying patients into clinical phenotypes, LS and non-LS, we identified additional genomic loci, underscoring distinct genetic architectures as we further characterize these clinical phenotypes.

Our findings suggest that the genetic factors influencing disease trajectory are highly specific, involving particular genomic loci, immune cell functions, and signaling pathways in genomic regions known to strongly influence immunoregulation, immune response, antigen presentation, and antigen receptor activity.

In chronic sarcoidosis, 12 independent genetic variants were identified, highlighting three genomic regions of importance, two of which are located in the HLA class II region on the major histocompatibility complex (MHC), encompassing *TSBP1, BTNL2,* and *HLA-DRB1* on chromosome 6, and *ANXA11, PLAC9, DYDC2,* and *TSPAN14* on chromosome 10.

In nonchronic sarcoidosis, we identified a large genomic region within the extensive MHC (chr6:25.1 Mb–33.1 Mb) containing 370 independent genetic variants and 205 genes associated with nonchronicity.

Further analyses, thereby stratifying disease trajectory by clinical phenotypes (LS and non-LS), yielded more focused findings on genomic loci implicated in disease trajectory.

SNP-heritability estimates indicated that common genome-wide SNPs explained a substantial proportion of variation in susceptibility to both chronic and nonchronic sarcoidosis. The estimates were numerically higher for nonchronic than chronic sarcoidosis, and the pattern repeated in LS and non-LS phenotypes. These findings suggest a stronger genetic influence on the nonchronic phenotype than on chronic sarcoidosis. However, additional analysis is needed because the intervals overlap.

Gene-based analyses identified genes linked to both chronic and nonchronic diseases, including those shared across both groups.

In chronic sarcoidosis, the observed associations with *HLA-DRA, HLA-DRB1, HLA-DQA1,* and *HLA-DPA1* genes support the hypothesis that chronic disease is driven by persistent antigen presentation, MHC class II–restricted immune activation, and immune regulation and dysregulation. Notably, these genes are core components of the HLA class II system, which presents antigen to CD4+ T cells [36]. Their predominance suggests that chronic sarcoidosis is strongly linked to ongoing antigen-driven adaptive immune activation, especially within the granulomatous inflammatory response [36]. Additional immune-regulatory genes, such as *BTNL2*, a gene well known for its association with sarcoidosis [37] and involvement in T-cell costimulation and regulation, suggest abnormal immune persistence as a contributing pathogenetic factor. Interestingly, *BTNL2* is often seen as a modifier of T-cell activation thresholds and has been shown to interact with γδ T cells to promote IL-17A production [38].

*TSPAN14* may affect immune signaling and membrane protein trafficking, as it has been reported to promote tumor growth by affecting immune function, platelet coagulation, angiogenesis, and infection [39]. *ANXA11* is a well-documented sarcoidosis-associated gene [40] and has key biological roles in membrane dynamics, apoptosis, and vesicle trafficking, potentially influencing antigen processing, cell survival, or inflammatory resolution. *ANXA11* has been suggested as a disease modulator in sarcoidosis [41–43], autoimmune diseases [44], and cancers [44, 45]. Other genes indicating transcriptional or epigenetic regulation include *RING1*, a component of the Polycomb repressive complex linked to epigenetic control of immune gene expression in chronic sarcoidosis, and *RXRB*, a nuclear receptor partner that can affect transcriptional activity, lipid metabolism, and immune responses. *PLAC9* suppresses cell growth by causing S-phase arrest through changes in cyclin and CDK expression and enhances cell motility, likely involving cyclins, E-cadherin, and vimentin [46]; it is also a key regulator of systemic metabolism, and it is reported to be associated with obesity [47, 48]. Little is known about *DYDC2* in sarcoidosis, but a recent study found that it is specifically expressed in ciliated epithelial cells, a subcluster of lung cells [49]. Ciliated epithelial cells are essential for maintaining the bronchial epithelium’s balance and integrity. The role of *TSPAN14* in sarcoidosis is unknown; however, accumulating evidence suggests this gene plays a role in non-small cell lung carcinoma (NSCLC) [50].

In nonchronic sarcoidosis, the gene-based approach identified a large set of extended MHC/HLA region-linked genes on 6p21, indicating broader immunological mechanisms and immune profiles. The biological implications of these genes suggest roles in antigen presentation, immune regulation, innate-adaptive crosstalk, and inflammatory control, rather than pointing to a single isolated pathway. Specifically, nonchronic sarcoidosis seems to be driven by a dense immune susceptibility area centered on: a) Class I HLA genes (*HLA-A, HLA-B, HLA-C, HLA-F, HLA-G*); b) Class II HLA genes (*HLA-DRA, DRB1, DQA1/2, DQB1/2, DPA1, DPB1, DMA/DMB, DOA/DOB*); c) Antigen processing machinery (*TAP1, TAP2, PSMB8, PSMB9*); and d) Immune regulatory genes (*BTNL2, TNF, LTA, LTB, NOTCH4, BRD2, CFB, C2, C4A, MICA, MICB, LST1, AIF1, NCR3, BAG6, RNF5,* and *TRIM* family genes).

Therefore, interactions among these genes likely affect disease development and resolution. Additionally, the extended-MHC immune framework centered on HLA class I and II antigen presentation, antigen-processing components, complement, cytokine genes, and immune-regulatory genes indicates a predominantly antigen-driven disease mechanism. This process involves both adaptive T-cell responses and innate immune surveillance, all under significant genetic regulation of immune recognition and the balance of inflammation.

Functional enrichment analyses based on publicly available transcriptome and epigenome datasets strengthened our findings by identifying significant genetic variants as molecular regulators that influence gene expression in relevant inflammation-related tissues and cell populations. Our focus on lung tissue, whole blood, and immune cells provided a more specific approach, leading to the identification of immune cells, lung tissue, and whole-blood eQTL polymorphisms (i.e., eQTL SNPs) for further exploration. These findings underscore the biological relevance of chronicity-associated SNPs, which align with gene expression patterns in relevant cells and tissues in sarcoidosis. Our eQTL SNP findings corroborate previously reported associations with sarcoidosis disease severity [51].

The chronic sarcoidosis pathway profile is consistent with a persistent antigen-driven granulomatous immune response characterized by strong MHC-mediated antigen presentation, T-cell receptor and co-stimulatory signaling, Th1 polarization, interferon-gamma activity, and macrophage activation. Enrichment of PD-1/PD-L1, CTLA4, IL-10, and T-cell exhaustion pathways suggests that chronic disease is accompanied by substantial immune regulatory feedback, reflecting prolonged immune stimulation rather than resolution.

Additional pathways involving dendritic-cell/NK-cell crosstalk, phagosome maturation, innate immune sensing, pyroptosis, and cellular stress responses indicate ongoing innate-adaptive immune interaction and chronic tissue remodeling. Overall, the data support a model of chronic sarcoidosis as a sustained, dysregulated Th1/macrophage-driven inflammatory state with active immune regulation and tissue stress.

Nonchronic sarcoidosis showed a highly concentrated genetic signal in the extended MHC region, with enrichment of HLA class I and class II genes, antigen-processing components (*TAP1, TAP2, PSMB8, PSMB9*), complement genes, and immune regulatory loci including *BTNL2, TNF, LTA, LTB, NOTCH4, and MICA/B*. This architecture supports a model in which nonchronic disease is driven by genetically determined antigen presentation and immune recognition, with coordinated involvement of adaptive T-cell responses, innate immune surveillance, complement activation, and regulatory mechanisms within the MHC locus. The accompanying chromatin- and transcription-related genes further suggest that epigenetic control of the HLA region may influence immune activation and disease expression.

### Limitations of the study

We defined disease trajectory into two phenotypes, chronic and nonchronic disease. Chronic disease spans a wide spectrum of severity, from patients with stable disease not requiring treatment to those with chronic active disease needing treatment; thus, we may have underestimated differences between chronic active and nonactive disease. In most studies, disease duration of more than two years in sarcoidosis has been defined as chronic. However, even if patients still have the disease after two years, they may recover later, or, conversely, they may have recovered at two years but later relapse.

Another limitation is that we may not be able to extrapolate to other ethnicities, and we have more chronic cases (usually 60-70% are reported to resolve), which we think is because New Karolinska Hospital is a referral center that attracts the most severe cases.

The study’s strengths include diagnosis validation conducted by pulmonologists, a homogeneous ethnic composition, a substantial sample size with long-term follow-up, and phenotypically well-characterized participants.

## Conclusions

Our investigation into disease trajectory in sarcoidosis identifies a shared immune-driven disease architecture, with both chronic and nonchronic phenotypes enriched for pathways involved in antigen presentation, T-cell activation, interferon signaling, and macrophage/dendritic cell function. However, chronic sarcoidosis showed stronger evidence of sustained adaptive immune activation, immune checkpoint signaling, and T-cell exhaustion. In contrast, nonchronic sarcoidosis exhibited broader extended-MHC genetic associations and pathways involving antigen processing and innate immune sensing. Genetic and eQTL analyses further underscore the importance of the extended MHC region, highlighting phenotype-specific immune regulatory molecules and networks that may influence disease persistence or resolution. Together, these findings support distinct immunogenetic mechanisms underlying sarcoidosis chronicity and provide a framework for future biomarker discovery and therapeutic development.

## Data Availability

The genomic data of individuals investigated in the European cohorts are not available due to GPDR regulations.

## Acknowledgments

Grants awarded to NVR include the Swedish Heart-Lung Foundation (No. 20200505 and 20200506). Grants awarded to SK include the Swedish Heart-Lung Foundation (No. 20250380, 20250710), the Swedish Research Council (No. 2022-00870), and Stockholm County Council (Grant No. 1001474). Support was also provided through the regional agreement on medical training and clinical research (ALF) between Stockholm County Council and Karolinska Institutet (Grant No. 998890). Grants awarded to LP include the Swedish National Research Council (Grant No. 2018-02884).

The authors thank all patients and the personnel involved in this study. We thank our research nurses, Margitha Dahl, Helene Blomqvist, and Susanne Schedin, for helping collect samples.

The computations and genomic data were enabled by resources provided by the National Academic Infrastructure for Supercomputing in Sweden (NAISS) and the Swedish National Infrastructure for Computing (SNIC) at the Uppsala Multidisciplinary Center for Advanced Computational Science (UPPMAX), partially funded by the Swedish Research Council through grant agreements no. 2022-06725 and no. 2018-05973.

## Ethics declarations

Conflict of interest: The authors declare no competing financial interests concerning the work described.

## Authors’ contributions

SJ carried out genetic and computational analyses. SK and PD characterized the Swedish cohort for clinical disease trajectory. LP and IK characterized the healthy controls in the Swedish cohort. OC contributed to the molecular profiling of the Swedish cohort. TI performed preliminary analyses and phenotype characterization. AE and AS helped establish and maintain the clinical Swedish cohort. MM, PS, and MS contributed to the clinical relevance and discussions on sarcoidosis disease trajectory. NVR led the study design and genetic analyses.

All authors participated equally in drafting and revising the manuscript.

## Notes

### Competing Interest Statement

The authors have declared no competing interest.

### Author Declarations

The ethics committee of the Stockholm Region Review Board gave ethical approval for this work (DNR: 2024-03916-02).

### Summary of Updates

The cohort was updated to increase the sample size. Additional genetic analyses, including subgroup analyses, were incorporated. Additional authors were included in recognition of their scientific contributions to the study.

